# *AutScore* – An integrative scoring approach for prioritization of ultra-rare autism spectrum disorder candidate variants from whole exome sequencing data

**DOI:** 10.1101/2024.01.24.24301544

**Authors:** Apurba Shil, Noa Arava, Noam Levi, Liron Levine, Hava Golan, Gal Meiri, Analya Michaelovski, Yair Tsadaka, Adi Aran, Idan Menashe

**Affiliations:** Department of Epidemiology, Biostatistics and Community Health Sciences, Faculty of Health Sciences, Ben-Gurion University of the Negev, Beer Sheva, Israel; Azrieli National Centre for Autism and Neurodevelopment Research, Ben-Gurion University of the Negev, Beer Sheva, Israel; Zlotowski Center for Neuroscience, Ben-Gurion University of the Negev, Beer Sheva, Israel; Bioinformatics Core Facility, Ben-Gurion University of the Negev, Beer-Sheva, Israel; Department of Physiology and Cell Biology, Faculty of Health Sciences, Ben-Gurion University of the Negev, Beer Sheva, Israel; Preschool Psychiatric Unit, Soroka University Medical Center, Beer Sheva, Israel; Child Development Center, Soroka University Medical Center, Beer Sheva, Israel; Child Development Center, Ministry of Health, Be’er Sheva 84100, Israel; Neuropediatric Unit, Shaare Zedek Medical Center, Jerusalem, Israel; Faculty of Medicine, The Hebrew University of Jerusalem, Jerusalem, Israel

**Keywords:** AutScore, candidate variants, ASD, WES, prioritization algorithm

## Abstract

**Background:** Discerning clinically relevant ASD candidate variants from whole-exome sequencing (WES) data is complex, time-consuming, and labor-intensive. To this end, we developed *AutScore*, an integrative prioritization algorithm of ASD candidate variants from WES data, and assessed its performance to detect clinically relevant variants.

**Methods:** We studied WES data from 581 ASD probands, and their parents registered in the Azrieli National Center database for Autism and Neurodevelopment Research. We focused on rare allele frequency <1%), high-quality proband-specific variants affecting genes associated with ASD or other neurodevelopmental disorders (NDDs). We assigned a score (i.e., *AutScore*) to each such variant based on their pathogenicity, clinical relevance, gene-disease association, and inheritance patterns. Finally, we compared the *AutScore* performance with the rating of clinical experts and the NDD variants prioritization algorithm, *AutoCasC*.

**Results:** Overall, 1161 ultra-rare variants distributed in 687 genes in 441 ASD probands were evaluated by *AutScore* with scores ranging from -4 to 25, with a mean ± SD of 5.89 ± 4.18. *AutScore* cut-off of ≥ 12 outperforms *AutoCasC* in detecting clinically relevant ASD variants, with a detection accuracy rate of 72.3% and an overall diagnostic yield of 11.9%. Sixteen variants with *AutScore* of ≥ 12 were distributed in fifteen novel ASD genes.

**Conclusion:** *AutScore* is an effective automated ranking system for ASD candidate variants that could be implemented in ASD clinical genetics pipelines.

## Introduction

Recent advances in high-throughput sequencing technologies have revolutionized genetic studies of complex diseases [1–7]. The emergence of next-generation sequencing (NGS) platforms has enabled genomic analyses at an unprecedented scale and resolution. These technologies have facilitated whole-genome sequencing (WGS) and whole-exome sequencing (WES) of large cohorts, unveiling novel disease-associated loci and providing deeper insights into the genetic architecture of complex disorders [1–9].

Detecting disease-causing variants from WES/WGS data is a complex task. Today, most clinical genetics labs that analyze WES/WGS data follow the American College of Medical Genetics and Genomics (ACMG) guidelines for interpreting sequence variants [10]. This mainly includes detecting high-quality variants with lower allele frequency and damaging effects on the protein function. Other factors usually considered are the segregation of the variant with the phenotype and existing evidence for the variant or gene association with the disease. To assist clinicians in this laborious process, several automated tools such as Exomiser [11], AMELIE [12], LIRICAL [13], AutoCasC [14], etc., have been devised to prioritize disease-specific variants (mainly single nucleotide variants [SNVs] and insertions/deletions [indels]) from WES/WGS data.

Autism Spectrum Disorder (ASD) is a complex neurodevelopmental disorder that has greatly benefited from the emergence of NGS technologies. Recent large-scale WES and WGS studies have identified thousands of ASD susceptibility genetic variants in hundreds of genes [5,15–20]. Nevertheless, despite these advances in ASD genetics, clinically meaningful genetic variants are identified only in 8% to 30% of affected probands [5,21,22]. Thus, there is a need for new approaches to facilitate the detection of ASD-specific variants from WES/WGS data. Here, we present an automated scoring approach called *AutScore* that integrates variant and gene-level information such as pathogenicity, deleteriousness, clinical relevance, gene-disease association, and gene-variant inheritance pattern from a wide range of bioinformatics tools and databases to generate a single score for prioritizing clinically relevant ASD candidate variants from WES data for simplex and multiplex families. We applied the *AutScore* to WES data from 581 Israeli ASD-affected probands and their parents. We assessed its performance by comparing the obtained results to a manual and blinded evaluation of the variants by clinicians and to *AutoCasC* [14], an existing variant prioritization tool for neurodevelopmental disorders (NDDs).

## Materials and Methods

### Study Sample

Our sample included 581 children diagnosed with ASD, registered with the Azrieli National Centre for Autism and Neurodevelopment Research (ANCAN) [23,24]. Based on clinical records, none of the parents had registered themselves with ASD, intellectual disability, or other neurodevelopmental disorders (NDDs). Genomic DNA was extracted from saliva samples from children and their parents using Oragene®•DNA (OG-500/575) collection kits (DNA Genotek, Canada).

### Whole Exome Sequencing (WES)

Whole Exome Sequencing (WES) analysis was conducted in two labs: (1) the Broad Institute as a part of the Autism Sequencing Consortium (ASC) project [25] and (2) the Clalit Health Services sequencing lab at Beilinson Hospital. WES was performed using Illumina HiSeq sequencers in both places, followed by the Illumina Nextera exome capture kit. The sequencing reads were aligned to human genome build 38 and aggregated into BAM/CRAM files. Then, the Genome Analysis Toolkit (GATK) [26] (Broad) or Illumina’s DRAGEN pipeline [27] (Beilinson) was used for variant discovery and the generation of joint variant calling format (vcf) files.

### Variant filtering and annotations

The multi-sample vcf files generated by the Genome Analysis Toolkit (GATK) and the DRAGEN platform were undertaken with identical procedures for variant filtering and annotation, as previously detailed [28]. Subsequently, we identified pathogenic (P), likely pathogenic (LP), or likely gene-disrupting (LGD) variants using the *InterVar* [29] tool in conjunction with our proprietary tool, *Psi-Variant* [28]. We kept only those LP/P/LGD variants that affected genes associated with ASD or other neurodevelopment disorders (NDDs) according to the SFARI gene [30] or the DisGeNET [31] databases for downstream analyses. Subsequently, 1161 candidate variants in 441 probands remained for further analysis (Fig. **1**).

**Fig. 1.**
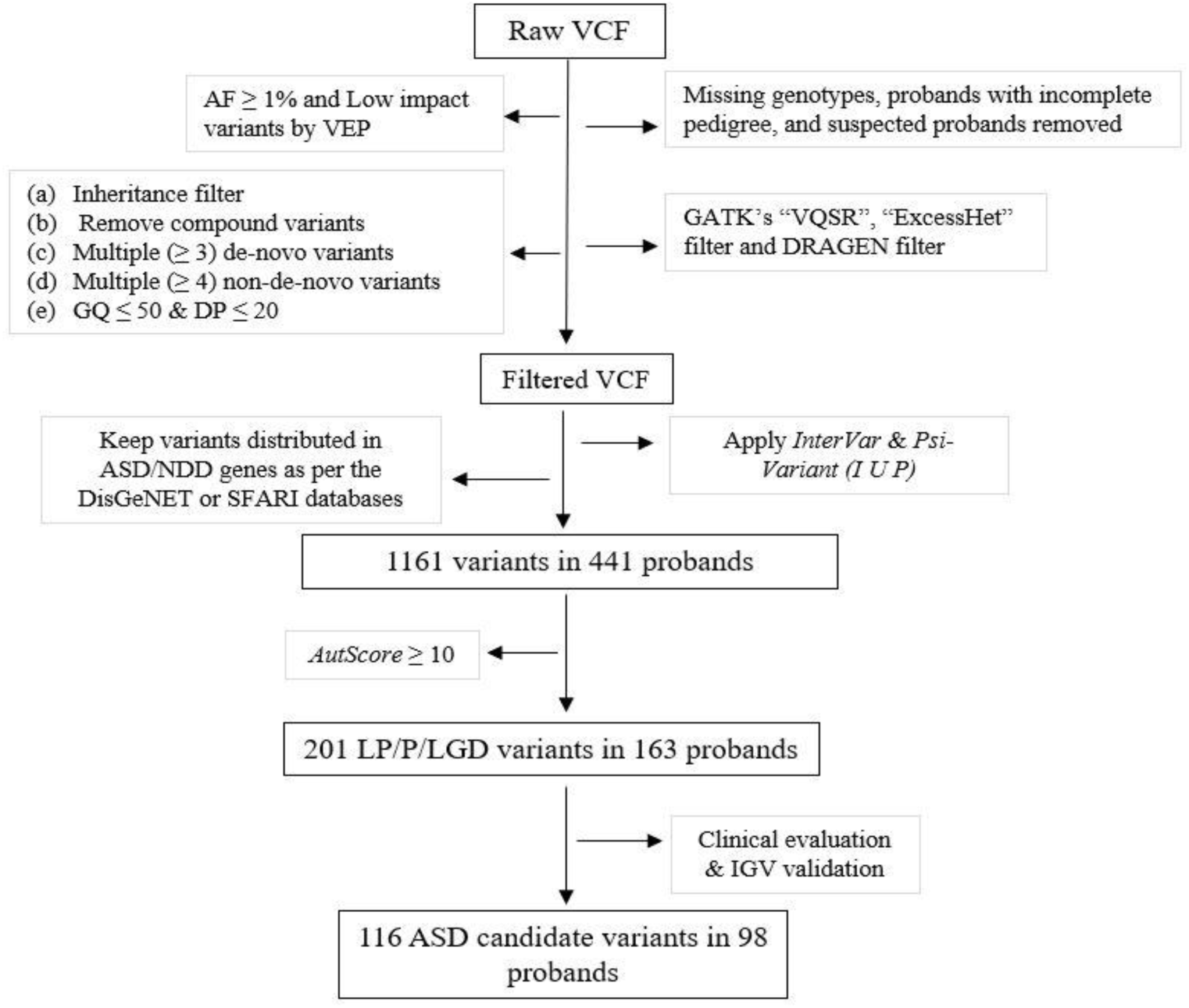
Analysis workflow for detecting ASD candidate variants from the WES data.

### Prioritization of ASD candidate variant

We developed a metric called *AutScore* to prioritize the detected list of ASD candidate variants as follows:

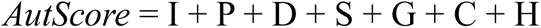

Where:

- I – indicates the pathogenicity of a variant based on *InterVar* [29] classification as follows: ‘benign’ = -3; ‘likely benign’ = -1; ‘variants of uncertain significance (VUS)’ = 0; ‘likely pathogenic’ = 3, and ‘pathogenic’ = 6.
- P – cumulatively assess the deleteriousness of a variant based on the following six in-silico tools (SIFT [32] (< 0.05), PolyPhen-2 [33] (≥ 0.15), CADD [34] (> 20), REVEL [35] (> 0.50), M_CAP [36] (> 0.025) and MPC [37] (≥ 2)). For each of these tools, a variant gets a score of 1 (deleterious) or 0 (benign), and these scores are aggregated to generate a single score ranging from 1 to 6.
- D – indicates the agreement of variant-phenotype segregation with the predicted segregation by the Domino tool [38] where agreement with Domino’s ‘very likely dominant/recessive’ classes = 2; agreement with Domino’s ‘likely dominant/recessive’ classes = 1; disagreement with Domino’s ‘very likely dominant/recessive’ classes = -2; disagreement with Domino’s ‘likely dominant/recessive’ classes = -1; and 0 were assigned for variants with Domino’s ‘either dominant or recessive’ segregation.
- S – indicated the strength of association of the affected gene with ASD according to the SFARI gene database [30] where ‘high confidence’ = 3; ‘strong candidate’ = 2; ‘suggestive evidence’ = 1; and not in SFARI database = 0.
- G – indicated the strength of association of the affected gene with ASD according to the DisGeNET database [31] where weak/no association (GDA=0 to 0.25) = 0: mild association (GDA=0.25 to 0.50) = 1: moderate association (GDA=0.50-0.75) = 2: strong association (GDA=0.75 and above) = 3.
- C – pathogenicity of a variant based on ClinVar [39] where ‘benign’ = -3; ‘likely benign’ = -1;’VUS’ = 0; ‘Likely pathogenic’ = 1; ‘Pathogenic’ = 3.
- H – segregation of variants in the family weighted as *(n^2^)-1* where n=number of probands in a family that carries the detected variants.

### Clinical genetics validation

Variants with *AutScore* ≥ 10 (top quartile of candidate variants scores) were visually validated using the IGV software [40] and then manually examined by clinical geneticists according to the standard ACMG/AMP guidelines [10]. The clinical experts assessed the likelihood of the variants contributing to the ASD phenotype of the child and assigned each variant one of the following rankings: ‘Likely,’ ‘Possibly,’ and ‘Unlikely’.

### Statistical Analysis

We used a Receiver Operating Characteristic (ROC) analysis to assess the performance of *AutScore* in detecting ASD candidate variants using the clinical experts’ rankings as the reference. We also accordingly compared the sensitivity, specificity, positive predictive value (PPV), negative predictive value (NPV), and accuracy. In addition, diagnostic yield (%) was computed as the proportion of the number of ASD probands that have at least one ASD candidate variant out of the total affected ASD probands that completed their WES analysis. We compared the performance of *AutScore* in detecting ASD candidate variants with the performance of *AutoCasC* [14], an existing variant prioritization tool for NDDs. The agreement between *AutScore* and *AutoCasC* scores, and between these scores and the clinical assessment ranking, were assessed using Pearson’s correlation and Cohen’s Kappa statistic, respectively.

### Software

Data storage, management, and analyses were conducted in a high-performing Linux cluster using Python version 3.5 and R version 1.1.456. All statistical analyses and data visualization were performed and incorporated into R.

## Results

A total of 1161 variants distributed in 687 genes in 441 ASD probands were evaluated by the *AutScore* algorithm. Variant’s scores ranged from -4 to 25, with a mean ± SD of 5.89 ± 4.18 (Fig. **2**). The clinical experts examined 201 (17.31%) variants with an *AutScore* of ≥ 10. Among these, 24 (11.9%) were suspected as false positive indels during the visual assessment using the IGV software and thus removed from subsequent analyses. Of the remaining 177 variants, 65 (36.7%) were ranked as ‘likely,’ 51 (28.8%) as ‘possibly,’ and 61 (34.5%) as ‘unlikely’ ASD candidate variants (Supplementary Table **S1**).

**Fig. 2.**
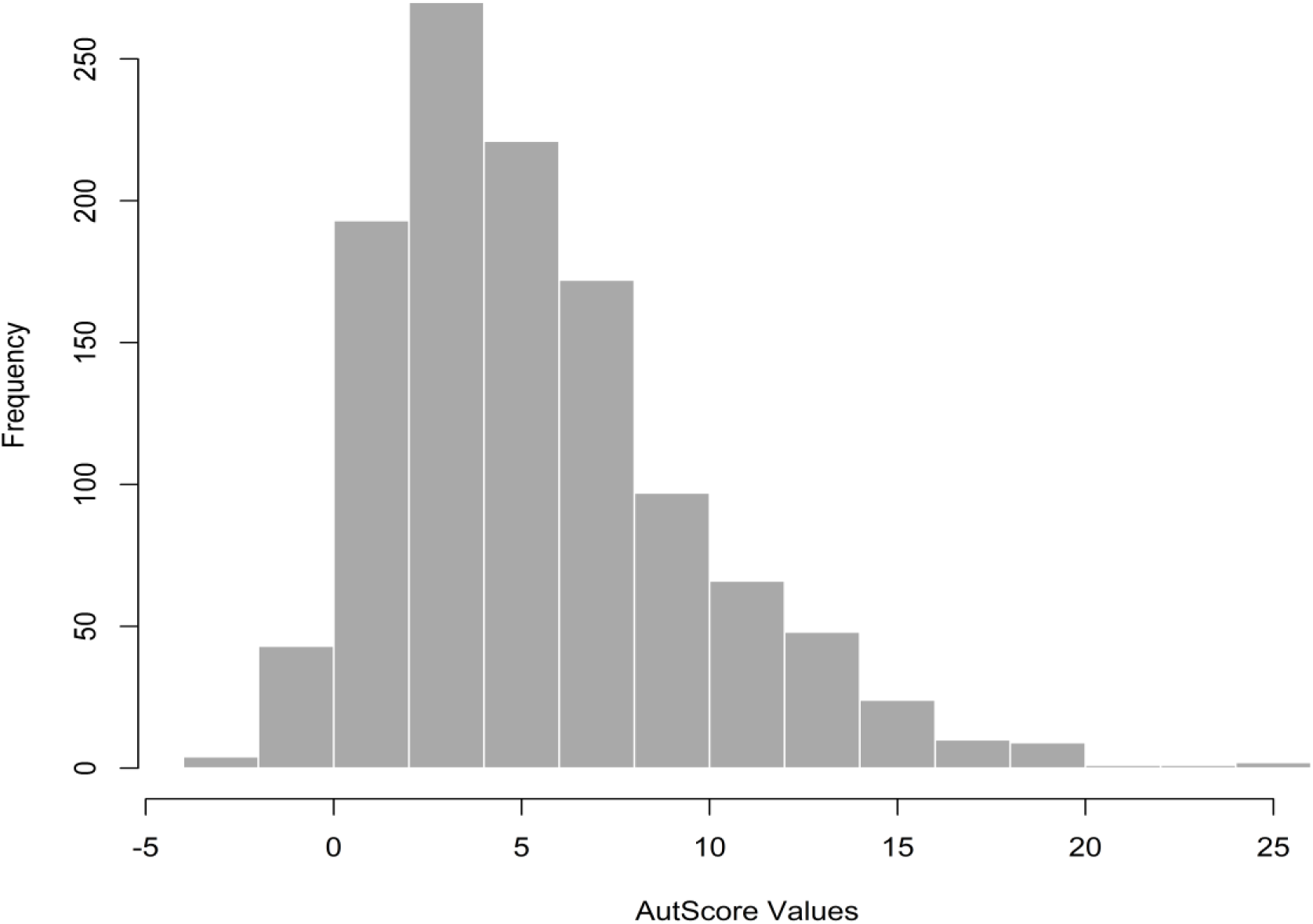
Histogram depicting the distribution of total LP/P/LGD variants assessed by *AutScore* (N=1161)

### Identifying an optimum *AutScore* cut-off

Two analyses were carried out to identify the optimal *AutScore* cut-off (Fig. **3**). First, an ROC analysis using the clinical experts’ ranking: “likely” as the true set of ASD candidate variants indicated that *AutScore* is an effective tool for detecting ASD clinically meaningful variants (AUC=0.843, 95% CI= 0.779-0.907) (Fig. **3A**). Applying Yuden J’s analysis to these data suggested that an *AutScore* of ≥ 12 would be the most effective cut-off (Yuden J=0.52). The same cut-off was also indicated by integrating detection accuracy and diagnostic yield (Max of Yield + Accuracy /10=17.04) (Fig. **3B**).

**Fig. 3.**
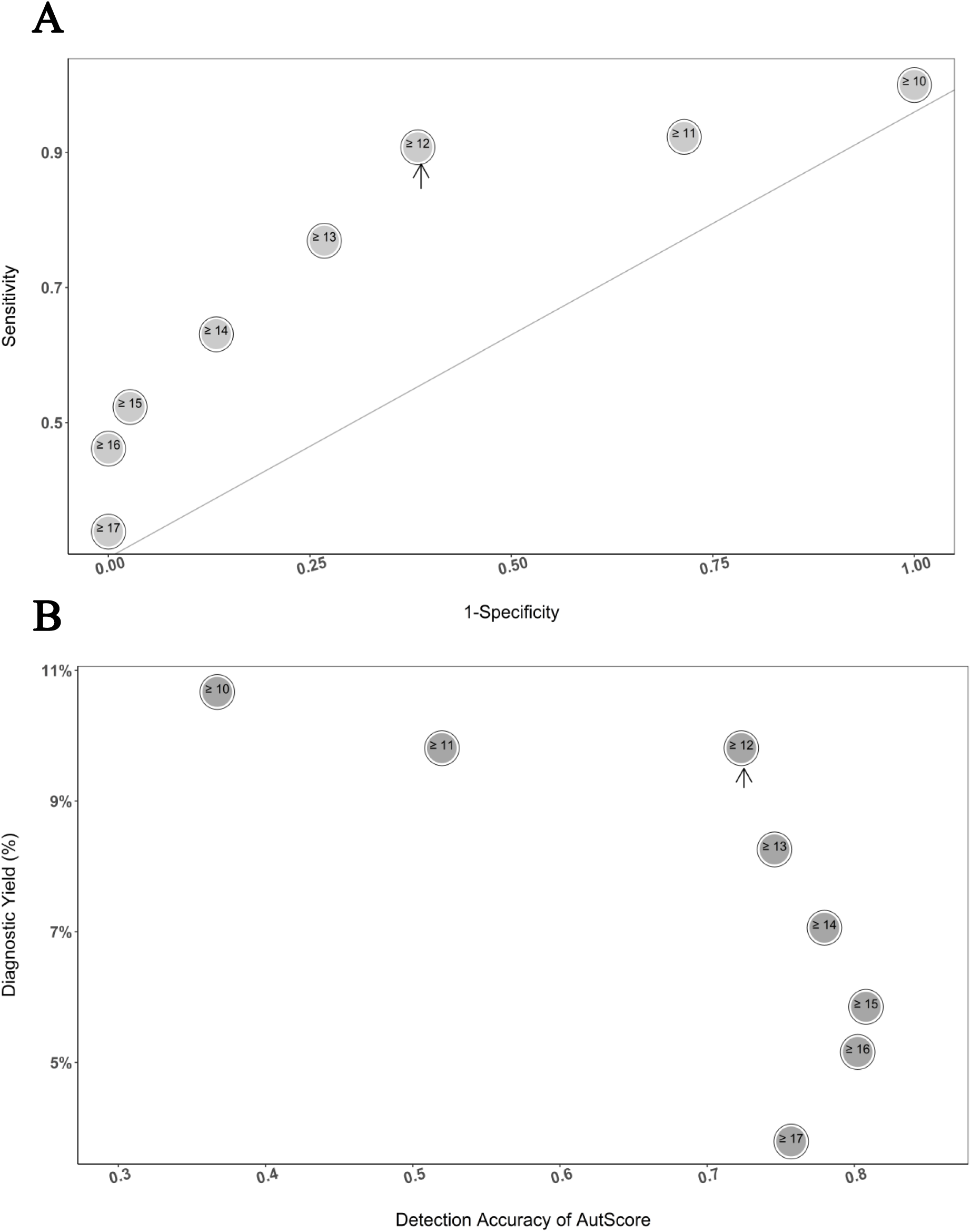
Assessing *AutScore’s* optimal cut-off for detection of ASD susceptibility variants. **A** A receiver operating characteristics (ROC) analysis for different *AutScore* cut-offs. An arrow indicates the best cut-off based on Yuden J’s statistics. **B** Scatterplot of the detection accuracy (x-axis) and the resulting diagnostic yield (Y-axis) for different *AutScore* cut-offs. An arrow indicates the best cut-off based on both values’ aggregated maximum.

### Comparing *AutScore* with *AutoCasC*

Next, we compared the performance of *AutScore* (using the selected cut-off ≥ 12) vis-à-vis the existing NDD prioritization tool, *AutoCasC*, using its recommended cut-off of >6 [14], in detecting ASD candidate variants (i.e., likely, possibly) (Fig. **4**). A moderate, but statistically significant correlation (r=0.58; p<0.05) was observed between *AutScore* and *AutoCasC*. Both tools had high sensitivity in detecting ASD variants using their recommended cut-off (0.91 and 0.92, respectively; Table **1**). Yet, *AutScore* outperformed in all other diagnostic characteristics except in its diagnostic yield (Specificity: 0.616, PPV: 0.578 and Accuracy: 72.3%; 95% C.I: 65.1%-78.8% vs. Specificity: 0.133, and PPV: 0.397 Accuracy: 43.5%; 95% C.I: 35.9%-51.3% respectively) (Table **1**). In addition, *AutScore* results had a better agreement with the clinical expert rankings than those of the *AutoCasC* (percentage agreement =72.3% and Cohen’s Kappa= 0.468 vs. percentage agreement=43.5% and Cohen’s Kappa= 0.04 respectively; Table **2**). The variant list (n=177) with *AutScore*, clinical assessments, and *AutoCasC* values is provided in Supplementary Table **S1**.

**Fig. 4.**
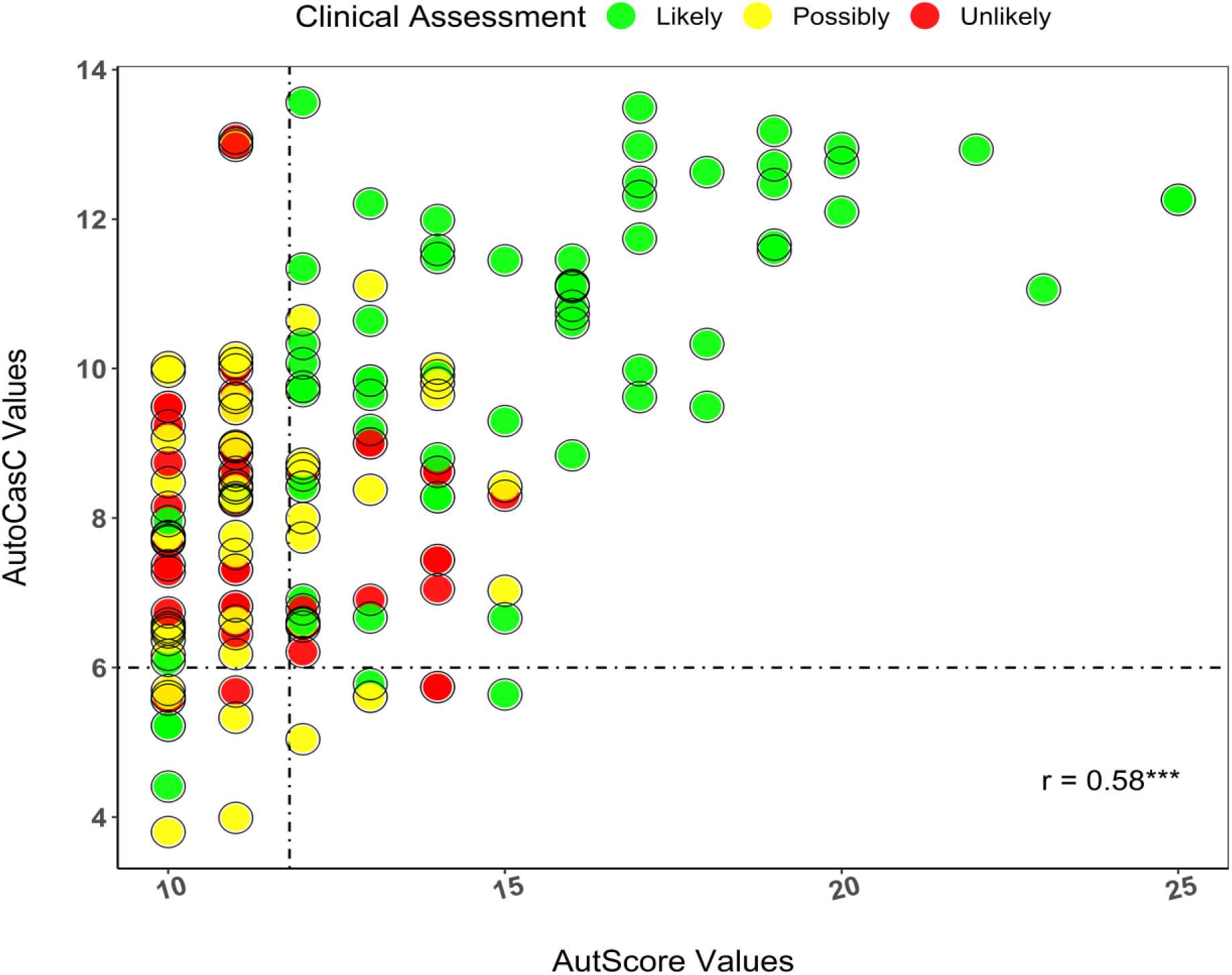
A clustered scatter diagram comparing the performance of *AutScore* (≥ 12), AutoCasC (> 6), and the clinical assessments (e.g., likely (green), possibly (yellow), and unlikely (red)).

**Table 1:** Comparing the performance between *AutScore (≥ 12)* and *AutoCasC (> 6)* in detecting ASD candidate variants.

| Scoring Approaches | Sensitivity | Specificity | PPV | Accuracy (95% C.I.) | Diagnostic Yield (Likely) | Diagnostic Yield (Likely+Possibly) | Yuden J |
| --- | --- | --- | --- | --- | --- | --- | --- |
| *AutScore* ($\geq 12$) | 0.91 | 0.62 | 0.58 | 0.72 (0.65, 0.79) | 9.81 | 11.9 | 0.52 |
| *AutoCasC* ($> 6$) | 0.92 | 0.13 | 0.40 | 0.43 (0.36, 0.51) | 9.98 | 15.5 | 0.06 |

**Table 2:** Concordance between *AutScore* (≥ 12), *AutoCasC* (> 6), and Clinical Expert Rankings in detecting ASD candidate variants (N=177 variants)

| Combinations | Percentage Agreement | Cohen's Kappa (P-Value) |
| --- | --- | --- |
| *AutScore* ($\geq 12$) Vs *AutoCasC* ($> 6$) | 57.6 | 0.07 (0.20) |
| *AutScore* ($\geq 12$) Vs Clinical Assessment | 72.3 | 0.45 (0.00) |
| Clinical Assessment Vs. *AutoCasC* ($> 6$) | 43.5 | 0.04 (0.26) |

### Characteristics of the LP/P/LGD variants detected by *AutScore*

Overall, 102 variants had an *AutScore* ≥12. Of these, 59, 18, and 25 variants were ranked as ‘likely’, ‘possibly’, and ‘unlikely’ ASD candidate variants, respectively, by the clinical experts (Table **3**). Most of the detected variants (45.1%) were distributed in high-confidence ASD genes according to the SFARI Gene database [30] (i.e., SFARI score of 1). Another 29 (28.4%) variants were detected in 23 genes (29.9%) not listed in the SFARI database and thus may be considered novel ASD genes. More than 90% of the detected variants were classified as LP/P according to the ACMG/AMP variant interpretation criteria [10], and more than 62% were denovo variants.

**Table 3:**
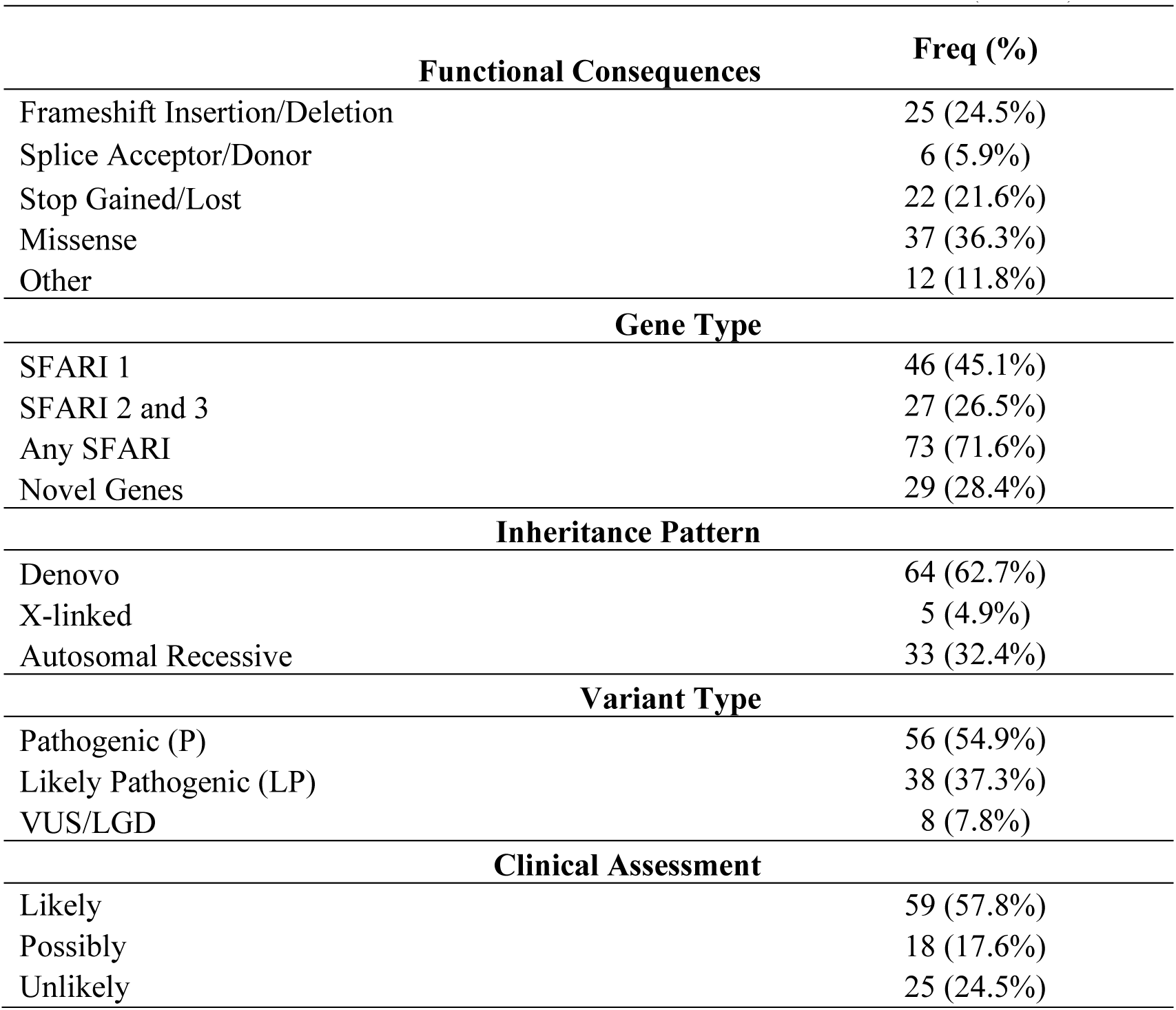
Characteristics of the detected variants with *AutScore* ≥ 12 (N=102)

## Discussion

Discerning clinically relevant ASD candidate variants from many variants poses a formidable challenge for clinical experts, demanding considerable time and effort. Here, we present *AutScore,* a novel bioinformatics prioritization tool that integrates variant and gene-level information to prioritize ASD candidate variants derived from WES data. *AutScore* can be integrated into an existing bioinformatic pipeline for WES data analysis by pre-installing the ACMG/AMP [10] variant interpretation tool InterVar [14] and our in-house tool *Psi-Variant* [28]. Although *AutScore* was initially designed to assess the ASD clinical relevance of rare autosomal SNVs, it can be adapted for analyses of copy number variants (CNVs), mitochondrial variants, and common heritable variants that are expected to enhance its applicability further.

Our results indicated that *AutScore* is highly efficient in detecting clinically relevant ASD variants. Using its most effective cut-off (i.e., ≥12), it achieves an overall diagnostic yield of 11.9%, comparable to results from prior studies [5,21,22]. We showed that *AutScore* outperforms the existing NDD variant prioritization tool, *AutoCasC* [14], in detecting clinically relevant ASD candidate variants. The higher accuracy of *AutScore* compared to *AutoCasC* is likely because it was explicitly designed to detect ASD candidate variants. At the same time, *AutoCasC* focuses on prioritizing candidate variants related to a broader range of NDDs.

The following limitations should be considered when using *AutScore*. First, the *AutScore* metric was established using a trial-and-error approach, assigning certain weights and penalties to its different elements. It is possible to mitigate this inherent subjectivity using a machine learning model-based prioritization score. Since such models require larger datasets of true ASD variants, we plan to upgrade to *AutScore* when such datasets are available. Second, *AutScore* is constrained to specific genes from the DisGeNET [31] and SFARI Gene [30] databases. Consequently, it might have missed some potential candidate variants in genes not cataloged in these databases. Third, the performance of *AutScore* data has not been assessed in WGS data. Hence, caution should be taken when applying this ranking tool to prioritize ASD candidate variants derived from WGS data. Fourth, the estimates derived from *AutScore,* including accuracy, PPV, and yield, were computed based on WES data from an ASD cohort within the Israeli population. Thus, these estimates could vary in other populations. Lastly, *AutScore* may not function optimally in cases involving probands with incomplete pedigree information and unknown segregation patterns.

## Conclusion

*AutScore* constitutes a highly effective automated ranking system designed to prioritize ASD candidate genetic variants in WES data. The utilization of *AutScore* holds the potential to significantly streamline the process of elucidating the specific genetic etiology of ASD within affected families. In doing so, it can contribute to expediting and enhancing the accuracy of clinical management and treatment strategies, ultimately leading to more effective interventions in the context of ASD.

## Supporting information

List of variants with AutScore values

## Data Availability

WES data were generated as part of the ASC and are available in dbGaP with study accession: phs000298.v4.p3. All the codes will be available upon reasonable request to the corresponding author.

## List of abbreviations

ASD: Autism Spectrum Disorder
SNVs: Single Nucleotide Variants
INDELs: Insertions/Deletions
LGD: Likely Gene Disrupting
LP/P/VUS: Likely Pathogenic/Pathogenic/Variants of Uncertain Significance
LoF: Loss of Function
CNVs: Copy Number Variants
WES: Whole Exome Sequencing
WGS: Whole Genome Sequencing
ACMG/AMP: American College of Medical Genetics and Genomics/Association of Molecular Pathology
GATK: Genome Analysis Toolkit
IQR: Interquartile Range
NDDs: Neurodevelopmental Disorders
PPV: Positive Predictive Value
NPV: Negative Predictive Value
SFARI: Simons Foundation Autism Research Initiative
OMIM: Online Mendelian Inheritance in Man
AUC: Area Under the Curve
ROC: Receiver Operating Characteristic

## Declarations

### Institutional Review Board Statement

The study was conducted according to the guidelines of the Declaration of Helsinki and approved by the Ethics Committee of Soroka University Medical Center (SOR-076-15; 17 April 2016).

### Ethics approval and consent to participate

Written consent was obtained from all parents of children involved in the study.

### Consent for publication

All the data from the registered families presented here are de-identified.

### Competing interests

The authors declare no competing interests.

### Funding

This study was funded by the Israel Science Foundation (#1092/21).

### Authors’ contributions

*Conceptualization*: A.S. and I.M.; *methodology*: A.S. and I.M.; *software*: A.S. and L.L.; *validation*: N.A. and N.L.; *formal analysis*: A.S.; *resources*: N.S., H.A.K, G.M., A.M., Y.T., A.A., H.G., and I.M.; *data curation*: A.S.; *writing—original draft preparation*: A.S. and I.M.; *writing—review and editing*: I.M., and A.S.; *supervision*: I.M.; *project administration*: I.M.; *funding acquisition*: I.M. All the authors have read and agreed to the published version of the manuscript.

## Acknowledgments

We thank the families who participated in this research; genetic studies would be impossible without their contributions.

Authors’ information (optional)

## Notes

### Competing Interest Statement

The authors have declared no competing interest.

### Summary of Updates

We have corrected Fig. 3B and Fig. 4.

